# Empowering Healthcare Professionals in West Africa □ A Feasibility Study and Qualitative Assessment of a Dietary Screening Tool to Identify Adults at High Risk of Hypertension

**DOI:** 10.1101/2023.11.09.23297914

**Authors:** Nimisoere P. Batubo, Nnenna M. Nwanze, Chizindu A. Alikor, Carolyn I. Auma, J. Bernadette Moore, Michael A. Zulyniak

**Affiliations:** Nutritional Epidemiology Group, School of Food Science and Nutrition, University of Leeds, Leeds, LS2 9JT; Department of Family Medicine, Rivers State University/Rivers State University of Teaching Hospitals, Port Harcourt, Rivers State, Nigeria; Department of Internal Medicine, Rivers State University/Rivers State University of Teaching Hospitals, Port Harcourt, Rivers State, Nigeria

**Keywords:** Food frequency questionnaire, feasibility study, hypertension risk, screening tool, West Africa

## Abstract

**Background:** Dietary risks significantly contribute to hypertension in Nigeria. Food frequency questionnaires (FFQs) can provide valuable dietary assessment but require rigorous validation and careful design to facilitate usability. This study assessed the feasibility and potential effectiveness of implementing a clinical screening tool for identifying adults at high risk of hypertension in West Africa.

**Materials and methods:** 58 consenting adult patients with hypertension and their caregivers and 35 healthcare professionals from a single-centre Nigerian hospital were recruited to complete a 27-item FFQ at two-time points and three 24-hour recalls for comparison in a mixed method study employing both quantitative questionnaires and qualitative techniques to elicit free form text. Data analyses were conducted using R software version 4.3.1 and NVivo version 14. The trial was registered with *ClinicalTrials.gov*: NCT05973760.

**Results:** The mean age of patients was 42.6 ± 11.9 years, with an average SBP of 140.3 ± 29.8 mmHg and a BMI of 29.5 ± 7.1 Kg/m^2^. The adherence rate was 87.9%, and the mean completion time was 7:37 minutes. 96.6% of patients found the FFQ easy to complete, comprehensive, and valuable. A minority reported difficulty (3.4%), discomfort (10.3%), and proposed additional foods (6.9%). Healthcare professionals considered the screening tool very important (82.9%) and expressed a willingness to adopt the tool, with some suggestions for clarification. Patients and healthcare professionals found the screening tool favourable for nutritional counselling in hypertension care.

**Conclusion:** The tailored screening tool (FFQ) demonstrated promising feasibility for integration into clinical care as assessed by patients and healthcare professionals. Successful implementation may benefit from proactive time management and addressing training needs. This user-centred approach provided key insights to refine FFQ and set the foundation for ongoing validity testing and evaluation in clinical practice.

## 1. Introduction

Hypertension, defined as sustained high blood pressure above systolic blood pressure ≥140 mm[Hg and diastolic blood pressure ≥90 mm[Hg, is the leading preventable risk factor for cardiovascular disease and the number one cause of death globally, responsible for over 10 million deaths annually (1). Approximately 40% of people aged 30-79 years have hypertension, with two-thirds of cases living in low- and middle-income countries, including African countries (WHO, 2021). Sub-Saharan Africa experiences a disproportionately high hypertension burden, with prevalence estimates ranging from 19% to 50% in adults (2, 3). The most recent data estimate a 17% increase in hypertension rates in Nigeria from 2010 to 2019 (4–6) and highlight that awareness, treatment, and control of hypertension remain suboptimal, with <30% of hypertensive adults in Nigerian managing to control their blood pressure (7, 8). This trend has been linked to poor healthcare access, high medication costs, and non-adherence to anti-hypertensive medications (9).

Unhealthy diets are a major modifiable risk factor for hypertension in Nigeria and globally (1, 10, 11). Our previous work reported that frequent intakes of diets high in salt, red meat, processed foods, fried foods, fat, and alcohol were associated with an increased likelihood of hypertension in Nigeria and other West African countries (12). In contrast, higher fruit and vegetable consumption appeared protective against hypertension. These findings provide region-specific evidence that unhealthy foods raise hypertension likelihood in West Africa. In 2018, the Nigerian Government estimated an average daily salt consumption of 10 grams/day among adults in Nigeria, which is approximately twice as high as WHO’s recommendation of <5 grams (13). Nigerian diets traditionally contain high sodium from added salt, bouillon cubes, salted-dried fish and salted-dried meat, with intakes increasing further from processed, restaurant, and fast foods (13–15). Therefore, optimising nutrition by encouraging restriction of sodium, processed and fried foods, and increased intake of fruits/vegetables, nuts/legumes, lean proteins, and healthy fats constitutes a vital component of hypertension prevention and treatment (9).

One of the key challenges in addressing hypertension in West African countries lies in the timely identification of individuals at high-risk (8). Limited tools for healthcare services, inadequate screening programs, and low awareness levels among the population contribute to delayed diagnosis and intervention (3, 16). In most cases, healthcare professionals are unable to capture a snapshot of the foods being consumed by their patients. Conversely, their patients are unaware that the foods they’re consuming are putting them at risk of hypertension until the condition progresses to a critical stage and they receive guidance.

Early identification of individuals at high risk of hypertension is paramount in mitigating its impact on public health (17). Healthcare professionals play a pivotal role in the early identification and management of hypertension (17). Their interactions with patients in clinical settings provide a crucial opportunity for early identification and intervention (3). However, the effectiveness of healthcare professionals in identifying individuals at high risk depends significantly on the availability of suitable screening tools that are both culturally sensitive and contextually appropriate (18). Therefore, timely intervention through accurate screening is critical for preventing the progression of chronic diseases and reducing associated morbidity and mortality rates (17, 19, 20).

Dietary assessment represents an essential first step for establishing the association between diet and disease and designing effective nutrition interventions for chronic conditions, including hypertension (21). Evidence suggests that culturally tailored food frequency questionnaires (FFQs) can improve validity and accuracy compared to non-specific tools (22–24). In West African countries, including Nigeria, cultural norms, dietary practices, and lifestyle patterns can influence the presentation and management of hypertension (25). Considering these challenges, it is imperative to develop and evaluate a screening tool specifically tailored to the cultural specificities of the African region. These tools should be acceptable, comprehensible, and non-burdensome for healthcare professionals and patients (18, 26).

The integration of validated screening tools into clinical care has the potential to augment disease risk assessment, improve patient counselling, and enhance treatment outcomes (27). Nevertheless, implementing screening tools within clinical settings has been constrained, primarily due to inadequate healthcare professional training, data interpretation challenges, lack of integration into electronic health records (EHRs) and clinical workflows, and time constraints (28, 29). To surmount these implementation barriers, it is imperative to ensure that (i) a study is designed to minimise the risk of bias and (ii) the tools and methods used are acceptable and appropriate for all participants (i.e., healthcare professionals and patients) before evaluating the efficacy of the tool itself. Therefore, this study aimed to assess the feasibility and opinions of a culturally tailored dietary screening tool in a clinical setting in Nigeria to screen individuals at risk of hypertension.

## 2. Materials and Methods

### 2.1. Study design and setting

This study utilised a single-centre, cross-sectional feasibility study with a mixed methods design combining quantitative and qualitative approaches to evaluate the acceptability, practicality, and perceived utility of implementing a screening tool for dietary assessment in routine clinical practice for hypertension management. The study was conducted in the outpatient (Family Medicine and Internal Medicine) clinics of Rivers State University Teaching Hospital, Port Harcourt, Rivers State, Nigeria. Patients interested in the study were referred directly by healthcare professionals to the study team or through recruitment fliers and posters. Eligibility was determined using a structured questionnaire (**Table 1**). Eligible patients were then allocated to either hypertension or non-hypertension categories. The study lasted for 4 weeks. We adhered to SPIRIT guidelines for reporting clinical trials (30).

**Table 1:** Inclusion and exclusion criteria.

| Inclusion criteria | Exclusion criteria |
| --- | --- |
| Age between 18 and 70 years<br>Men and women<br>Hypertensive or non-hypertensive individual<br>Individuals who have been residents in Nigeria for the past 2 years.<br>Ability to read, write, and communicate over the phone in English<br>Individuals who gave their consent to participate. | Individuals < 18 years or > 70 years of age<br>Pregnant women or intend to become pregnant or breastfeeding woman<br>Diagnosis of other chronic diseases such as cancer, diabetes, renal failure, endocrine diseases, and previous and recent history of cardiovascular disease (CVD) and stroke.<br>Individuals on dietary restriction or recent changes to their diet or food<br>Individuals who are currently enrolled in other studies |

### 2.2. Participant recruitment

The patients were recruited during routine clinic visits over 4 weeks between June and July 2023 through recruitment posters displayed in key areas of the hospital, clinician referrals, and engaging discussions during morning briefing sessions where vital signs are taken from patients in the internal medicine and family medicine departments of RSUTH. The healthcare professionals identified eligible participants based on study criteria and informational posters and referred interested participants to the study team for further screening. In total, 90 patients expressed interest in the study and were screened for eligibility, which yielded 66 patients. Additionally, 35 healthcare professionals were recruited through informational files distributed in the hospital and presentations at clinic meetings. The healthcare professionals gave written informed consent to provide qualitative feedback on the screening tools.

### 2.3. Ethics approval and informed consent

The study protocol was submitted to the following ethics boards. Business, Earth & Environment, Social Sciences (AREA FREC) Committee, University of Leeds, Leeds, United Kingdom on 21^st^ March 2023, and the Rivers State University Teaching Hospital Research Ethics Committee in Port Harcourt, Nigeria on 20^th^ March 2023 and granted final approval with the approval number: 0484 on 28/04/2023 and approval number: RSUTH/REC/2023316 on 30/03/2023 respectively. All participants provided informed consent to participate in this research study. The trial was registered at clinicaltrials.gov (Trial Registration: NCT05973760).

### 2.4. Eligible participants

Eligible participants were adults between the ages of 18-70 years, including both men and women, who were resident in Nigeria for at least two years and could read, write, and communicate in English. Both hypertensive and non-hypertensive individuals were eligible, provided they did not have dietary restrictions or recent diet changes, were free from cancer, diabetes, renal failure, endocrine disorders, previous cardiovascular disease, or stroke, and were not enrolled in other studies. The full inclusion and exclusion criteria are presented in **Table 1**. Before enrolment, each participant read a simplified version of the participant information sheet and was provided an opportunity to ask questions of the study staff to ensure informed consent to participate in the study.

### 2.5. Study protocol

The study was a single-cantered feasibility study conducted among adult outpatients (Family Medicine and Internal Medicine) clinics of the Rivers State University Teaching Hospital in Nigeria between July 2023 and August 2023. Participants were seen at a screening visit 1-4 weeks to assess eligibility per established inclusion/exclusion criteria before the dietary assessment (**Table 1**). If eligibility criteria were met, the patients were allocated into hypertensive and normotensive groups based on their history of hypertension, and informed consent was obtained from participants. The feasibility of the screening tool was evaluated in a small number of participants (*n*=101) consisting of 66 patients and 35 healthcare professionals (**Fig 1**).

**Fig 1.**
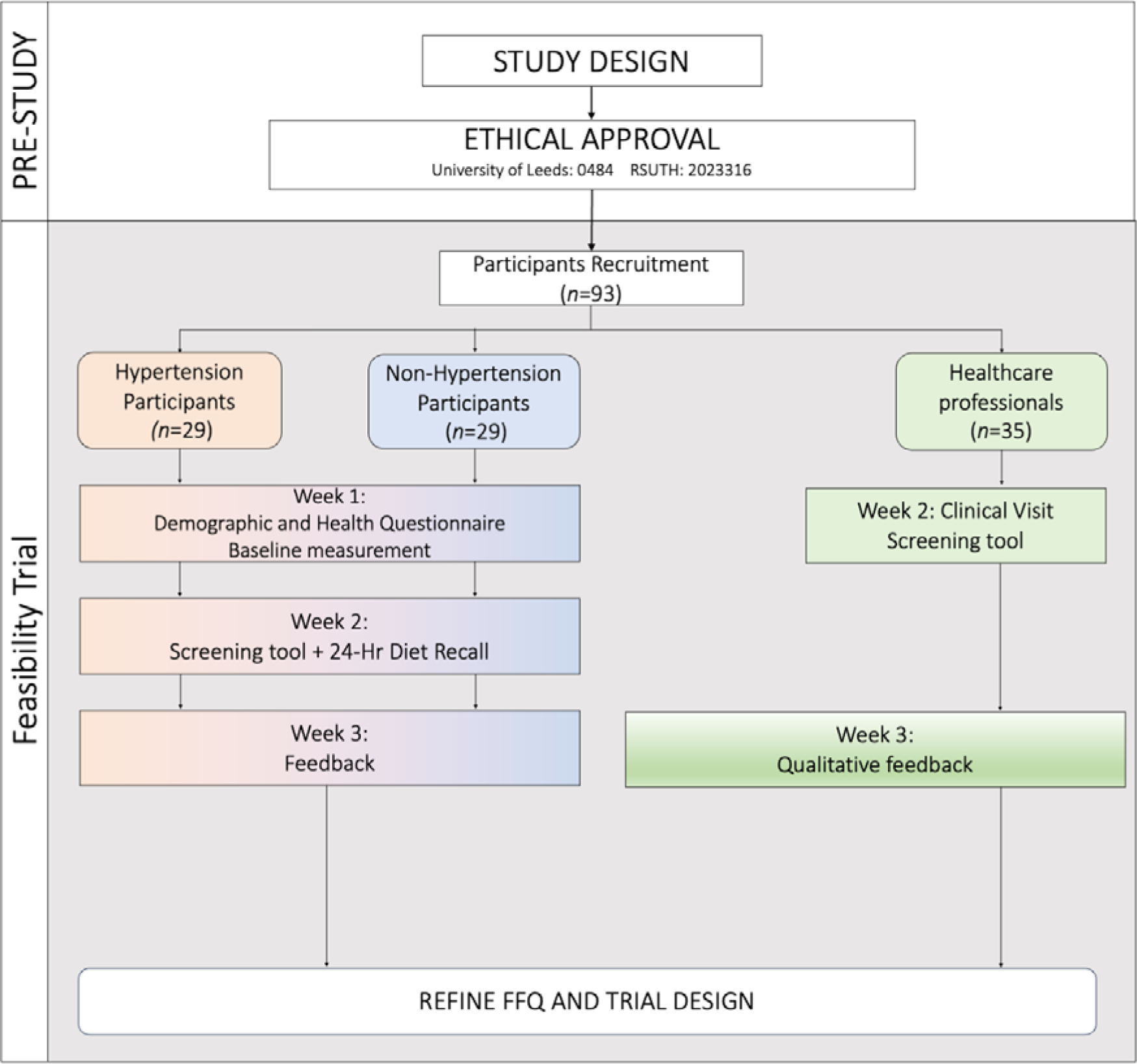
Participant selection and sequence of assessments flowchart. FFQ: food frequency questionnaire, 24 HR: 24-hour dietary recalls, BP: Blood pressure, H: Height, W: Weight.

In the week 1 post-allocation study visit, participants underwent baseline assessments consisting of demographic and health questionnaires, anthropometric measurements (height and weight), and blood pressure evaluation. The participant’s height and body weight were measured two times using a standard stadiometer (model number: DG2301, China). The body mass index (BMI) was calculated from the measurement of the height and body weight of each patient. The participant’s blood pressure was measured two times in the non-dominant arm using an automated mercury sphygmomanometer (model number: ZK-BB68, Shenzhen, China).

On the second study visit, eligible patients and healthcare professionals filled out the screening tools (27-item semi-quantitative food frequency questionnaire) (**S1 Table**). Alongside the screening tool, the first 24-hour dietary recall (24HR) was used to garner dietary intake data from the patients. Finally, on the third study visit, eligible patients and healthcare professionals completed a one-time survey examining the completion rates, clarity of questions, ease of use, cultural appropriateness and difficulty encountered while answering the questions. Additionally, the healthcare professionals provide qualitative feedback on the clinical relevance, their perceptions surrounding the integration of the screening tool into clinical practice and the potential impact of the screening tool on patient care in Nigerian healthcare settings and suggest improvements to optimise the questionnaire’s effectiveness for the prevention and dietary management of hypertension. The healthcare professional’s survey was adapted from previous studies on clinical adoption of health technologies (31). Questions utilised a 5-point Likert-type scale and open-ended formats.

### 2.6. The screening tool (FFQ)

The screening tool for this study was a semi-quantitative food frequency questionnaire (FFQ) containing 27 food groups covering common Nigerian foods and dishes informed by our previous systematic review and meta-analysis on dietary factors associated with hypertension in West Africa (12) and the Nigerian national nutrition guidelines on non-communicable disease prevention (32) (**S1 Table**). For each food group, participants reported consumption frequency ‘on a typical week’ over the past month or so, and answers corresponded to one out of four options, with categories ranging from ‘rarely or never, ‘1-2 times/week’, ‘3-5 times/week’, ‘daily’, and ‘more than once per day’. Additional questions related to salt intake behaviour were appended to the FFQ. The FFQ was self-administered. According to the Flesch-Kincaid scale, the reading level was at a 6th-grade level (33).

### 2.7. Participant compensation

Patients and healthcare professionals received a gift card incentive of £5 (approximately LJ6,000) after completing the third study visit involving the FFQ and surveys. Healthcare professionals received a gift card incentive of £5 (LJ6,000) upon completing the one-time feasibility and perceptions survey about the screening tool.

### 2.8. Sample size

The target sample size was 50 patients allocated into hypertensive and non-hypertensive groups, with a ratio of 1:1 between groups. This was estimated based on published recommendations for feasibility studies and qualitative research to allow for sufficient power for the validation analyses and achieving thematic saturation for qualitative feedback (34–37), (38) and was adjusted to account for up to a 20% dropout rate (39, 40). For healthcare professionals providing qualitative feedback, a sample of 25 was deemed adequate.

### 2.9. Statistical analysis

The quantitative and qualitative analyses were conducted in R version 4.3.1 (41) and NVivo version 14 (42), respectively. The quantitative data were exported to R statistical software and analysed using descriptive statistics, including frequencies, means, and standard deviations. Results of paired t-tests indicated whether, on average, there were differences between men and women. Completion rates, retention and adherence were analysed as percentages. The mean completion time between the first and second visits was calculated. Patient and healthcare professionals’ responses to 5-point Likert scale questions were analysed as means and standard deviations.

Qualitative responses from the healthcare professionals underwent thematic analysis to identify common themes through an iterative process and stepwise process of open coding, categorisation, and consensus building among analysts (43, 44). First, the qualitative feedback from healthcare professionals, which was collected through open-ended questionnaires and interviews, transcribed verbatim, was entered into Microsoft Excel for organisation by participant group. The responses were then imported into NVivo 14 qualitative analysis software (42) for thematic analysis. This followed an iterative process, which involved initial open-coding to extract broad themes, focused-coding to categorise responses under identified themes, constant comparison between groups, and saturation assessment. Two independent coders analysed the data to enhance rigour. Through consensus building, themes and sub-themes related to feasibility, usability, cultural appropriateness, limitations, and integration considerations emerged. The themes were quantified by calculating the percentage of participants mentioning each theme. Representative quotes were identified. The results were reported as mean ± SD and median for continuous data and *n* (%) for categorical data, and *p*-values <0.05 were considered statistically significant.

## 3. Results

### 3.1. Patient’s feasibility outcomes

#### 3.1.1. Patient’s characteristics

Ninety patients were initially screened; 66 met the inclusion criteria, provided informed consent, and enrolled in the study. Twenty-four patients were excluded (10 were on dietary restriction, 6 had a recent change of diet, and 8 were aged less than 18 years). After exclusions, 58 patients completed the study, including 40 women (69%) and 18 men (31%). Eight patients were excluded due to incomplete FFQ data (**Fig 2**). The mean age was 42.6 ± 11.9 years, with males averaging 40.5 ± 12.1 years and females 43.5 ± 11.8 years. Educational levels ranged from primary schooling (3.4%) to postgraduate degrees (24.1%). Regarding marital status, 67.2% were married, 31.0% were single, and 1.7% were widowed. Employment displayed diversity, with 24.1% self-employed and 37.9% in government positions. Over half (55.2%) reported a family history of hypertension, indicating a potential inherited predisposition. Additionally, 50% had hypertension, varying from under one year (31.0%) to over five years (41.4%). Only 22 (75.9%) of the 29 patients with hypertension were on antihypertensive medications (**Table 2**).

**Fig 2.**
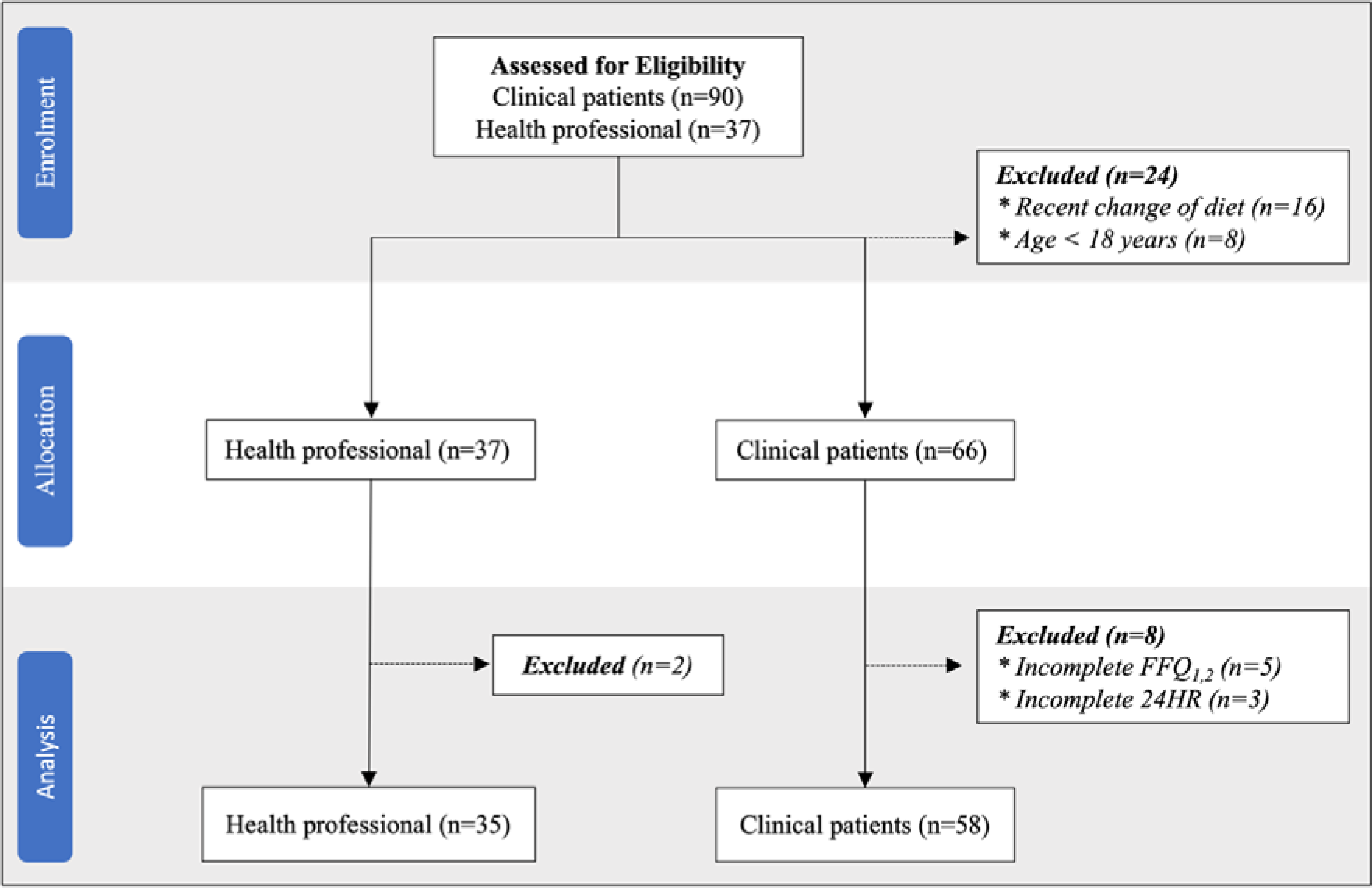
Patient recruitment and enrolment flowchart. FFQ: Food frequency questionnaire, 24HR: 24-hour dietary recall, n: number

**Table 2:** Sociodemographic characteristics of patients (*n*=58)

| Characteristics | Overall | Men | Women |
| --- | --- | --- | --- |
| Age (years), mean ± SD | 42.6 ± 11.9 | 40.5 ± 12.1 | 43.5 ± 11.8 |
| Sex, n (%) | 58 (100) | 18 (31.0) | 40 (69.0) |
| Education n (%) |  |  |  |
| Primary | 2 (3.4) | 0 | 2 (3.40) |
| Secondary | 12 (20.7) | 2 (3.45) | 10 (17.20) |
| High school | 4 (6.9) | 2 (3.45) | 2 (3.45) |
| University | 26 (44.8) | 9 (15.50) | 17 (29.30) |
| Postgraduate | 14 (24.1) | 5 (8.62) | 9 (15.50) |
| Marital status, n (%) |  |  |  |
| Married | 39 (67.2) | 10 (17.2) | 29 (50.0) |
| Single | 18 (31.0) | 8 (13.8) | 10 (17.2) |
| Widow | 1 (1.7) | 0 | 1 (1.7) |
| Employment, n (%) |  |  |  |
| Homemaker | 3 (5.2) | 0 | 3 (5.2) |
| Student | 3 (5.2) | 1 (1.7) | 2 (3.5) |
| Unemployed | 3 (5.2) | 0 | 3 (5.2) |
| None-paid | 1 (1.7) | 0 | 1 (1.7) |
| Self-employed | 14 (24.1) | 2 (3.5) | 12 (20.7) |
| Non-governmental | 9 (15.5) | 3 (5.2) | 6 (10.3) |
| Government | 22 (37.9) | 10 (17.2) | 12 (20.7) |
| Retired | 3 (5.2) | 1 (1.7) | 2 (3.5) |
| Family history of Hypertension, n (%) |  |  |  |
| No | 26 (44.8) | 10 (17.2) | 16 (27.6) |
| Yes | 32 (55.2) | 8 (13.8) | 24 (41.4) |
| Hypertension n (%) |  |  |  |
| No | 29 (50.0) | 9 (15.5) | 20 (34.5) |
| Yes | 29 (50.0) | 9 (15.5) | 20 (34.5) |
| Years of hypertension n (%) |  |  |  |
| < 1 year | 9 (31.0) | 3 (5.2) | 7 (12.1) |
| 1-5 years | 8 (27.6) | 2 (3.5) | 5 (8.6) |
| > 5 years | 12 (41.4) | 3 (5.2) | 9 (15.5) |
| Antihypertensive medications use n (%) |  |  |  |
| No | 7 (24.1) | 2 (6.9) | 5 (17.2) |
| Yes | 22 (75.9) | 6 (20.7) | 16 (55.2) |

Data are presented as: *n*= frequency, %= percentage, mean ± SD= Standard deviation

**Table 3**. presents the cross-sectional measurements of the patients. On average, the patients had a body weight of 79.4 ± 17.2 kg. Men weighed more, with an average of 87.3 ±19.0 kilograms, compared to women, who averaged 75.8 ± 15.2 kilograms. The overall height averaged 1.65 ± 0.1 meters, with males being taller at 1.77 ± 0.1 meters as opposed to females, whose height averaged at 1.60 ± 0.1 meters. The mean Body mass index (BMI) calculated was 29.5 ± 7.1 kg/m^2^. The average systolic and diastolic blood pressure were 140.3 ± 22.9 mmHg and 87.4 ± 17.3 mmHg, respectively, across all patients. Male patients had slightly higher mean systolic (143.3 ± 24.7mmHg) and diastolic (90.5 ± 19.6 mmHg) pressures compared to female patients (138.9 ± 23.8 mmHg and 86.0 ± 16.2 mmHg, respectively).

**Table 3:** Cross-sectional measurements of patients (*n*=58)

| Characteristics | Overall | Men | Women | p-value |
| --- | --- | --- | --- | --- |
| Height (m) | $1.65 \pm 0.1$ | $1.77 \pm 0.1$ | $1.60 \pm 0.1$ | <0.001 |
| Body weight (kg) | $79.4 \pm 17.2$ | $87.3 \pm 19.0$ | $75.8 \pm 15.2$ | 0.032 |
| Blood pressure |  |  |  |  |
| SBP (mmHg) | $140.3 \pm 29.8$ | $143.3 \pm 24.7$ | $138.9 \pm 23.8$ | 0.525 |
| DBP (mmHg) | $87.4 \pm 17.3$ | $90.5 \pm 19.6$ | $86.0 \pm 16.2$ | 0.397 |
| Body mass index (kg/m<sup>2</sup>) | $29.5 \pm 7.1$ | $28.2 \pm 5.9$ | $29.6 \pm 7.5$ | 0.364 |
| Completion Time |  |  |  |  |
| Mean | $7.37 \pm 2.28$ | | | |
| First visit | $7.71 \pm 2.51$ | $7.83 \pm 2.71$ | $7.65 \pm 2.46$ | 0.808 |
| Second visit | $7.03 \pm 2.03$ | $7.33 \pm 1.64$ | $6.90 \pm 2.18$ | 0.408 |

Data are presented as: mean ± SD=standard deviation
p-value between men and women at $p < 0.05$

#### 3.1.2. Retention

Of the 66 enrolled patients, 58 completed the screening tools (FFQ) during the initial and follow-up study visits, with a retention rate of 87.9%, demonstrating a high level of participant engagement and commitment throughout the study duration (**Fig 2**).

#### 3.1.3. Adherence

In evaluating participant adherence, the study demonstrated a high level of compliance. Out of the 66 patients initially recruited, 58 completed all required assessments and activities, resulting in an adherence rate of approximately 87.9% (**Fig 2**). Notably, eight participants faced challenges in adhering to the study protocols, resulting in instances of non-adherence. Specifically, three participants completed the initial Food Frequency Questionnaire (FFQ) but could not proceed with the second FFQ due to work schedule constraints. Additionally, five participants completed the first 24-hour recall but encountered challenges in completing the subsequent recalls, with reasons including poor network signal (*n*=3) and participants not responding (*n*=5).

#### 3.1.4. Completion rates and time

The completion rate for the 27-item FFQ was 100% (66 patients) at the first study visit. At three weeks, the completion rate was 87.9% (58/66 patients) (**Fig 2**). The mean time to complete the FFQ was 7.71 ± 2.51 minutes at the first study visit and 7.03 ± 2.03 minutes at three weeks (**Table 3**). Time to complete decreased over time as patients became more familiar with the process.

#### 3.1.5. Patients feedback

Patient’s perspectives on FFQ usability and efficacy are provided in **Table 4**. Most patients (96.6%) found the FFQ clear, appropriate, and easy to use. However, a minority (3.4%) reported some questions as unclear, especially on ‘more than once a day’ and salt intake frequency options (‘never/rarely’, ‘Sometimes’, ‘Usually and ‘Always’). Additionally, 10.3% expressed sensitivity to certain questions on topics like fruit intake and salt use behaviours. However, most (82.8%) were comfortable with the FFQ. For example, regarding fruit intake, one participant stated:

**Table 4.** Patients feedback on the screening tool (FFQ)

| Feedback responses | Yes | No |
| --- | --- | --- |
|  | *n* (%) | *n* (%) |
| Clarity, appropriate, and easy to use | 56 (96.6) | 2 (3.4) |
| Unclear or difficult questions | 6 (10.3) | 52 (89.7) |
| Discomfort and sensitivity in questions | 10 (17.2) | 48 (82.8) |
| Missing foods or beverages | 4 (6.9) | 54 (93.1) |
| Technical difficulties and factors influencing responses | 1 (1.7) | 57 (98.3) |
| Comprehensive assessment of food intake | 56 (96.6) | 2 (3.4) |
| Usefulness for clinicians | 56 (96.6) | 2 (3.4) |

Data are presented as: *n*= frequency, %= percentage

> “I feel uncomfortable because I don’t normally eat fruit and vegetables because of financial constraints” regarding fruit intake” (female, 40-45 years, >5 years hypertensive).

On salt use, another shared:

> “Most of my meals I bought from a food vendor, and I don’t know that it is important to check the amount of salt in food labels” (male, 30-35 years, non-hypertensive).

Regarding comprehensive food coverage, 6.9% noted missing items like unripe plantain, corn flour, Tofu (soybeans), and Ukwa (breadfruit). Just 1.7% reported technical difficulties influencing responses. Notably, 96.6% of participants believed that the questionnaire provided a comprehensive assessment of their food intake, highlighting its effectiveness, and an equally high percentage (96.6%) considered it useful for healthcare professionals in providing personalised nutritional support to patients.<colcnt=2>

### 3.2. Healthcare professional’s feasibility outcomes

#### 3.2.1. Healthcare professional’s characteristics

In total, 37 healthcare professionals enrolled in the feasibility survey of the FFQ. Thirty-five completed the survey. Two were excluded due to incomplete data (**Fig 2**). Years of experience ranged from 3 years to 20 years in practice.

#### 3.2.2. Perceived importance and current practices

The survey results demonstrated the importance of dietary assessment among healthcare professionals for effective hypertension management (**Table 5**). All healthcare professionals (100%) considered dietary assessment as ’very important or important’, underscoring its recognition as a crucial component in hypertension prevention and care. 71.4% of healthcare professionals reported conducting a routine dietary assessment in clinical practice occasionally as part of hypertension management; however, a smaller proportion of healthcare professionals conduct dietary assessments monthly (11.4%) or weekly evaluations (11.4%), while only a minority (5.7%) never performed assessments.

**Table 5:** Healthcare professional’s feedback on the feasibility of integrating the FFQ in clinical practice.

| S/N | Variables | Category | n (%) |
| --- | --- | --- | --- |
| 1 | Importance of dietary assessment | Very important | 29 (82.9) |
|  |  | Important | 6 (17.1) |
| 2 | Dietary assessment in practices | Monthly | 4 (11.4) |
|  |  | Never | 2 (5.7) |
|  |  | Occasionally | 25 (71.4) |
|  |  | Weekly | 4 (11.4) |
| 3 | Confidence in utilizing dietary data | Confident | 19 (54.3) |
|  |  | Very confident | 8 (22.9) |
|  |  | Neutral | 6 (17.1) |
|  |  | Not sure | 2 (5.7) |
| 4 | Challenges in assessing food intake | No | 17 (48.6) |
|  |  | Yes | 18 (51.4) |
| 5 | Existence of culturally-specific tools | No specific tools | 13 (37.1) |
|  |  | Yes, but not culturally specific | 7 (20.0) |
|  |  | Not sure | 14 (40.0) |
|  |  | Yes, culturally specific tools are available | 1 (2.9) |
| 6 | Clarity of questions | Yes | 34 (97.1) |
|  |  | No | 1 (2.9) |
| 7 | Difficulty of questions | No | 29 (82.9) |
|  |  | Yes | 6 (17.1) |
| 8 | Relevance of the new dietary tool (FFQ) | Highly relevant | 15 (42.9) |
|  |  | Relevant | 18 (51.4) |
|  |  | Neutral | 1 (2.9) |
|  |  | Not sure | 1 (2.9) |
| 9 | How valuable is the new tool (FFQ) | Very valuable | 20 (57.1) |
|  |  | Valuable | 14 (40.0) |
|  |  | Neutral | 1 (2.9) |

Data are presented as: n= frequency, %= percentage

#### 3.2.3. Current practices and confidence in tool usage

The survey assessed the confidence level among healthcare professionals in interpreting and applying dietary data for decisions regarding hypertension care, as outlined in **Table 5**. Most (77.2%) expressed ‘confident or very confident’. However, a notable portion, 17.1%, expressed a neutral stance, while 5.7% conveyed uncertainty. These findings suggest various confidence levels among healthcare professionals in utilising dietary data. Furthermore, over half (51.4%) encountered difficulties analysing patients’ dietary intake to determine hypertension risks and make dietary recommendations. This highlights the complexity involved in existing dietary assessment tools. In contrast, 48.6% did not encounter such challenges, indicating a balanced distribution of experiences among healthcare professionals. When evaluating healthcare professionals’ awareness of culturally specific dietary screening or assessment tools tailored for hypertension in Nigeria, a significant majority (57.1%) indicated either the absence of such tools or noted that available tools lacked cultural specificity. Meanwhile, 40% were uncertain about the existence of any dietary assessment tools. Only a small minority (2.9%) answered that culturally specific tools exist.

#### 3.2.4. Clarity and user-friendliness of the FFQ

The results from questions examining the perceived clarity of the new dietary assessment tool are presented in **Table 5**. An overwhelming majority (97.1%) of healthcare professionals found the questions easy to understand and straightforward, indicating a high level of clarity in the questionnaire. Only a small minority (2.9%) reported difficulty, specifically with questions about salt intake behaviour, suggesting an overall positive response regarding comprehensibility. Additionally, the survey assessed whether any questions posed challenges or were unclear for participants (**Table 5**). Once again, a majority (82.9%) reported no issues, underscoring the high level of clarity and user-friendliness in the questionnaire. However, a minority (17.1%) found some questions unclear.

#### 3.2.5. Perceived utility of the dietary assessment tool

In assessing the healthcare professional’s perceived relevance of the dietary tool for identifying and addressing dietary factors contributing to hypertension among patients as outlined in **Table 5**. Most healthcare professionals found it relevant (51.4%) or highly relevant (42.9%). Some were uncertain (2.9%) or neutral (2.9%). Overall, the results underscore the perceived potential of the tool for effectively targeting dietary factors associated with hypertension. Regarding the perceived value of the tool’s information for formulating individualised dietary recommendations for patients, most healthcare professionals (97.1%) deemed it valuable (**Table 5**). Only a small minority expressed a neutral stance (2.9%) (**Table 5**). This indicates a consensus among healthcare professionals regarding the high value of the data obtained through the tool for tailoring dietary advice to individual patients.

#### 3.2.6. Perceived feasibility and acceptability

The perception of the healthcare professionals on the feasibility and acceptability of integrating the newly introduced dietary assessment tool, the Food Frequency Questionnaire (FFQ), into their clinical workflows and patient consultations is detailed in **Table 6**. Among the healthcare professionals, a significant majority (82.9%) expressed confidence in the feasibility of integrating the tool (FFQ) into routine clinical practice. However, a small proportion (11.4%) were neutral or unfeasible (2.9%). Furthermore, in assessing the potential impacts on workflow, most healthcare professionals (85.7%) believed the integration would be feasible without disrupting established workflows. However, some uncertainty existed, with 14.3% expressing uncertainty about potential impacts and 2.9% asserting that it would be infeasible without affecting workflow. These findings collectively demonstrate an optimistic outlook on the feasibility of integrating the FFQ into the routine workflow of healthcare professionals.

**Table 6:** Perceived implementation and challenges of the new tool.

| S/N | Variables | Category | n (%) |
| --- | --- | --- | --- |
| 1 | Perceived feasibility of integration | No | 1 (2.9) |
|  |  | Not sure | 5 (14.3) |
|  |  | Yes | 29 (82.9) |
| 2 | Impact on workflow | Feasible | 24 (68.6) |
|  |  | Highly feasible | 6 (17.1) |
|  |  | Neutral | 4 (11.4) |
|  |  | Not feasible | 1 (2.9) |
| 3 | Foreseeable challenges in integration | No | 17 (48.6) |
|  |  | Not sure | 10 (28.6) |
|  |  | Yes | 8 (22.9) |
| 4 | Likelihood of implementation | Very likely | 17 (48.6) |
|  |  | Likely | 16 (45.7) |
|  |  | Neutral | 1 (2.9) |
|  |  | Very unlikely | 1 (2.9) |

Data are presented as: n= frequency, %= percentage

Moreover, the survey assessed the potential challenges that healthcare professionals foresaw in integrating the FFQ into their clinical practice (**Table 6**). Encouragingly, nearly half of the healthcare professionals (48.6%) believed there would be no discernible challenges, indicating an overall positive outlook. However, a significant portion (28.6%) expressed uncertainty, highlighting the need for comprehensive training and support programs to address potential concerns. Additionally, 22.9% of respondents anticipated challenges, underscoring the importance of proactive measures to identify and mitigate potential obstacles during the integration process.

In terms of the likelihood of successful implementation of the tool in routine clinical practice in Nigeria, the majority (94.3%) of healthcare professionals demonstrated a high degree of confidence, asserting that the implementation of the FFQ for personalised dietary counselling and assessment would ‘very likely or likely’, reflecting substantial confidence in the tool’s potential to enhance patient outcomes and contribute meaningfully to hypertension management. Only a marginal proportion (2.9%) maintained a neutral stance, indicating a need for targeted efforts to address any reservations or uncertainties. Similarly, only 2.9% deemed success to be ‘very unlikely’, further affirming the overall confidence in the potential benefits of the FFQ (**Table 6**). 94.3% of healthcare professionals indicated that the new tool is ‘likely or very likely’ to be implemented for personalised dietary counselling and evaluation. This reflects substantial confidence in the tool’s potential to improve patient outcomes and meaningfully contribute to hypertension management. Only 2.9% of healthcare professionals were neutral or deemed success ‘very unlikely’.

#### 3.2.7. Healthcare professional’s feedback

The qualitative feedback provided by the healthcare professionals yielded six themes that emerged from the content analysis of fragments and statements. These themes encompassed a comprehensive exploration of various aspects pertaining to screening tools, including knowledge of dietary screening tools, the effectiveness of existing screening tools, challenges in assessing dietary habits, essential features of a dietary screening tool, perceived benefits and challenges of integration of the tool, and perceived implementation and suggestions.

##### Knowledge of dietary screening tools

This theme, ‘knowledge of dietary screening tools,’ assessed the healthcare familiarity with alternative tailored dietary assessment tools and interventions for hypertension management. The healthcare professionals (97.1%) predominantly conveyed a lack of prior engagement with alternative dietary assessment tools (**Table 5**). Their responses consistently indicated a dearth of previous experience or utilisation of such tools, with many explicitly stating “no” or indicating a lack of familiarity. One healthcare professional stated:

> “I haven’t really used any specific dietary tools before. It’s quite new to me, to be honest” (male, 40-45 years, clinician).

Notably, 94.3% of healthcare professionals expressed interest in the new screening tool (FFQ), underscoring its perceived potential benefits, particularly its cultural adaptability (**Table 5**). Additionally, some healthcare professionals provided constructive feedback, suggesting improvements such as a more comprehensive representation of foods from northern Nigeria and enhanced functionality that does not rely on internet access. In this regard, one healthcare professional remarked:

> “I find the new screening tool quite intriguing. It seems adaptable to our cultural context, which is really important. However, it would be great to see more foods from the northern region of Nigeria included” (female, 40-45 years, clinician).

##### Effectiveness of existing assessment tools

The majority of healthcare professionals (97.1%) indicated a paucity or unfamiliarity with culturally tailored dietary tools for capturing indigenous foods and difficulties in quantifying some specific diets in cultural meals (**Table 5**). Furthermore, some healthcare professionals emphasised the need for a simplified tool in the form of a pamphlet or checklists with culturally adapted foods that can effectively address dietary assessment:

> “I find that the existing dietary tools don’t really consider our cultural context. There’s a gap when it comes to addressing indigenous foods, which are a significant part of our diet. Plus, quantifying the amount of salt and other diets in our cultural dishes using these tools can be quite challenging” (female, 40-45 years, clinician).
>
> “I firmly advocate for culturally tailored dietary tools. Take Awadu, for instance; it’s not even accounted for in any existing dietary assessment tool. Additionally, we must give due consideration to culturally specific food groups in our dietary assessments” (female, 46-50 years, clinician).

##### Challenges in assessing dietary habits

Almost half of the healthcare professionals (48.6%) expressed obstacles in evaluating and analysing patients’ food intake for hypertension risk assessment and subsequent dietary recommendations. These challenges encompass difficulties in accurately quantifying culturally specific meals, unfamiliar indigenous foods, concerns about recall bias, a lack of tailored assessment tools, and patients’ limited capacity to measure specific components of their food precisely. Additionally, healthcare professionals raised issues related to patient education and motivation alongside cultural beliefs, posing additional challenges in the intake of dietary data from patients:

> “In my experience, quantifying food content can be a real challenge. There are times when it’s quite tricky to measure, and patients often struggle to accurately estimate the proportions of salt, fats and seasonings used in their cultural meals” (female, 40-45 years, clinician).
>
> “I find myself unfamiliar with the specific foods that patients mention when assessing their food intake. It’s an area where I feel a bit out of my depth in clinical practice” (male, 40-45 years, clinician).

##### Essential features of a dietary screening tool

The healthcare professionals highlighted essential features they would consider crucial in a dietary screening tool for effective integration into clinical practice for assessing dietary intake in hypertension prevention and management. Firstly, they emphasised that the tool should be culturally tailored to the specific population and contain common foods in relation to specific health conditions. Secondly, the tool should be easy and quick to use, accessible, and able to capture intake and quantify processed food consumption, such as fried food or takeaway meals, fibre, salt intake, and saturated fat intake. Lastly, the healthcare professionals emphasised the tool should consider lifestyle and cultural factors that influence food choices and noted that language barriers should be addressed:

> “I believe a short dietary assessment tool should cover common food, be easy to use, short and culturally adaptable, and able to quantifiable content and consideration of portion sizes are also vital for accurate assessment” (male, 40-45 years, clinician).

##### Perceived benefits and challenges of integration of the new tool

A substantial majority, exceeding 90% of healthcare professionals, perceived the new screening tool as a relevant and valuable tool for assessing the food intake in patients and evaluating hypertension risk (**Table 5**). The integration of this tool into hypertension prevention and management has been identified by healthcare professionals as feasible (82.9%) and would not negatively impact workflow, conferring several benefits (**Table 7**). Nevertheless, they also recognised a spectrum of possible challenges that could be associated with its integration into clinical practice in hypertension management in Nigeria. Top on the list of perceived challenges was time constraints, especially in a busy clinic, where healthcare professionals and patients may be impatient due to limited consultation time, as illustrated in **Table 7**. In this regard, one healthcare professional stated:

**Table 7:** Benefits and challenges of implementing the screening tool.

| S/N | Benefits | Challenges |
| --- | --- | --- |
| 1 | Promotes healthy lifestyles and provides education on various food types and their benefits for individuals with hypertension. | Time constraints in busy outpatient settings may lead to increased waiting times during consultations. |
| 2 | Facilitates early identification of individuals at risk of hypertension, contributing to prevention through risk factor reduction. | Lack of patient awareness about the tool's existence and availability of the tool in patient folders may not always be ensured. |
| 3 | Enables healthcare professionals to offer tailored dietary advice, aligning recommendations with each patient's specific needs and preferences. | Effectiveness relies on patients providing accurate dietary information, which may not always occur due to memory or reporting inaccuracies. |
| 4 | Supports ongoing assessment and tracking of dietary components influencing hypertension, enabling timely detection of potential concerns | Limited comprehensive research on the effects of all foods on hypertension may make it challenging to provide tailored recommendations. |
| 5 | Assures holistic care for hypertension patients, providing detailed information about meal patterns for more effective management. | Insufficient manpower may affect tool implementation. Accessibility to trained personnel knowledgeable about local foods may pose challenges. |
| 6 | Encourages adoption of lifestyle practices beneficial for hypertension management and fosters a patient-centered approach to care. | Language barriers and patient literacy may hinder effective tool utilization, impacting its overall effectiveness. |
| 7 | Enhances speed and quality of patient care, streamlining risk assessment and progress monitoring in nutrition therapy. | Concerns about accurately measuring food, salt, and fat/oil intake, a critical factor in hypertension management. |

> In our busy clinic, both clinicians and patients can be quite impatient, especially during peak hours. Filling out the form might be rushed, and there’s a risk that the dietary assessment could be skipped (male, 40-45 years, clinician).

##### Perceived implementation and suggestions

In addition to identifying potential challenges of integration of the new tool, healthcare professionals offered insightful suggestions to facilitate the seamless integration of the dietary assessment tool into clinical practice. They advocated for targeted advocacy efforts, specifically directed towards the Ministry of Health and other health organisations, to secure support and endorsement for the implementation of the tool. Additionally, effective time management during consultations was deemed crucial, prompting suggestions such as condensing the questionnaire or streamlining the assessment process and creating a clear and simplified questionnaire, ensuring comprehensibility for individuals with varying levels of education. Language considerations were highlighted, with recommendations to translate the tool into Nigeria’s three major languages to enhance accessibility and acceptance among patients. Some healthcare professionals also proposed developing the tool into a software application, a progressive step towards improved accessibility and usability for both healthcare professionals and patients.

> “I feel a streamlined questionnaire, with clear language, is vital for everyone at different education levels and translating the tool into the three major languages in Nigeria will enhance its accessibility among the patient population” (male, 40-45 years, clinician).

## 4. Discussion

The present study explored the feasibility of integrating a validated dietary screening tool (Food Frequency Questionnaire) into clinical practice for dietary assessment and prevention of hypertension in Nigeria. This feasibility study provides important preliminary data on the potential adoption and utilisation of a tailored 27-item food frequency questionnaire (FFQ) for dietary assessment to prevent and manage hypertension in Nigeria. The results indicate an overall positive reception of the tool among patients and healthcare professionals. The high completion rates, satisfaction, and perceived utility support the acceptability and promise of the tool.

### 4.1. Acceptability and user engagement

The high initial completion rate of 100% and 87.9% at three weeks far exceeds rates observed in similar feasibility studies involving FFQs, which typically ranged from 70-90% (45–47). This high level of engagement indicates a strong acceptance among patients and aligns with another study with an 85% rate in a 2018 web based FFQ validation trial (47). Furthermore, the high retention rate of 88% compares favourably to prior FFQ trials, which retained 75-85% of patients enrolled in the study (48), demonstrating a high level of acceptability of the screening tools. According to a review by Hebert et al., retention rates above 80% are considered robust for dietary assessment studies, signifying a solid commitment from the patients for the studies (49). Therefore, the high retention rate achieved in this study favourably compares to previous feasibility studies involving FFQ, which retained 75-85% of participants (50). These collective findings suggest a strong acceptance and compliance with the feasibility study protocol.

The average time taken to complete the trial, approximately < 8 minutes (7.37 minutes), aligns with the goal of developing an efficient tool, which is consistent with the 5-15 minutes range reported in previous studies focusing on the feasibility and validation of FFQ (47, 48, 51, 52). Kristal et al. emphasised the importance of short completion times, defining them as ≤15 minutes in order to minimise participant burden (50). The slight reduction from 7.42 to 7.02 minutes at the second administration suggests that patients find the process undemanding and that it can be completed more quickly once it is familiar. In many healthcare facilities across Nigeria, patients typically experience hospital waiting times ranging from 83.7 to 144 minutes with a mean pre-consultation interval of 48.7 minutes (53–55). Therefore, across most healthcare settings in Nigeria, the implementation of this screening tool, which requires less than 8 minutes for completion, is unlikely to disrupt clinical workflow. The overwhelmingly positive feedback from patients, with 96.6% expressing satisfaction, mirrors the result observed in FFQ feasibility studies reporting satisfaction rates ranging from 70-90% (56). This high satisfaction indicates a positive user experience with an easily understandable, user-friendly format.

### 4.2. Clinical need and perceived utility

The consensus among healthcare professionals regarding the significance of dietary assessment in hypertension management is consistent with well-established evidence on the association between dietary factors and blood pressure (12, 57-60). However, the reported infrequency of assessments in routine practice supports the need for tailored tools. Previous studies also highlight a gap between recommended lifestyle counselling and actual clinical practice implementation (61–64). The anticipated benefits from the new screening tool, including education, early risk detection, personalised guidance, monitoring, and patient management, align with the multifaceted utility of such a tool. The study by Kristal et al. further emphasised the importance of healthcare professionals’ confidence in FFQ data for effective counselling, which is a pivotal factor in the clinical adoption (50). Therefore, the positive perceptions of utility among this sample of healthcare professionals are promising. Additionally, the fact that 97.1% of healthcare professionals regarded the screening tool (FFQ) as valuable for providing individualised dietary recommendations further affirms its perceived clinical relevance.

### 4.3. Cultural appropriateness

The common view among healthcare professionals that existing dietary assessment tools or methods lack cultural specificity substantiates existing conclusions that current approaches often inadequately capture the dietary habits of ethnic minority populations (65–68). The healthcare professionals emphasised the critical need to account for indigenous foods and quantify components, aligning with previous studies that demonstrated how traditional FFQs frequently misrepresent the dietary intake of ethnic minority groups (24, 69, 70). Thus, there is an evident niche for culturally tailored tools like the FFQ that this tool will evaluate in the follow-up study.

### 4.4. Implementation considerations

Over half of healthcare professionals expressed full confidence in utilising FFQ data, a factor previously identified by Kristal et al. as facilitating implementation (50). However, enhancement through training could promote broader confidence. Supporting healthcare professionals in applying FFQ insights may be warranted based on requests for guidance on counselling applications. While healthcare professionals acknowledged time constraints as a challenge, most believed implementation was feasible without causing significant disruption to workflow, aligning with studies demonstrating minimal time burdens associated with FFQ administration (27, 69).

Almost half of healthcare professionals foresaw no significant implementation challenges, reflecting a sense of optimism. However, some uncertainty remained regarding potential barriers related to patient factors, such as health literacy. Several reviews have highlighted the importance of user-centred design and a careful introduction, emphasising the influence of patient acceptance on the success of the FFQ (52, 71-73). Thus, a participatory implementation approach that engages all stakeholders could prove beneficial for the successful integration and implementation of the FFQ into clinical practice in Nigeria (74–77).

### 4.5. Strengths and Limitations

This study provides promising evidence supporting the FFQ as an acceptable, useful, and culturally appropriate tool for the dietary assessment and prevention of hypertension in Nigeria. This study demonstrates a comprehensive approach by employing both quantitative metrics and qualitative feedback, providing a multifaceted assessment of the feasibility of the FFQ from the perspectives of both patients and healthcare professionals. The pragmatic clinical environment enhances the applicability of the findings to the real-world context of hypertension care in Nigeria. The examination of numerous metrics provides comprehensive insights into retention, adherence, completion, relevance, acceptability, and anticipated integration barriers. Importantly, gathering user-centred feedback from providers allowed for evidence-based refinements that address their specific needs and priorities. The convergence of quantitative and qualitative findings offers complementary perspectives, enriching the overall feasibility evaluation.

Nonetheless, some limitations warrant acknowledgement. The single-centre design restricts generalizability to other Nigerian regions and clinical settings. Unlike past research, no comparative FFQ analysis occurred. Additionally, this study assessed perceived feasibility rather than actual pre-post implementation challenges. While potential barriers to integration were anticipated, the intricate details of real-world workflow integration remain underexplored. Furthermore, only perception-based, rather than objective data on clinical utility and patient outcomes, was captured. Finally, a longitudinal follow-up to assess whether acceptability endures over time was not conducted.

### 4.6. Relevance to clinical practice

The findings of this feasibility study bear important implications for the potential integration of the FFQ as a dietary assessment tool within clinical practice. The high completion rates, coupled with positive user feedback, suggest a promising acceptance of the tool among patients and healthcare professionals. Moreover, the minimal time investment required for FFQ completion aligns with the practical demands of a clinical setting. The identified challenges and recommendations provided by healthcare professionals offer valuable insights for refining the tool’s implementation process. These considerations are pivotal in guiding future steps towards the effective incorporation of the screening tool (FFQ) into routine clinical practice, ensuring its seamless integration and utility in the dietary assessment and prevention of hypertension among the Nigerian patient population.

## Conclusions

This study provides initial promising evidence to support the screening tool (FFQ) as an acceptable, useful, and culturally appropriate dietary assessment tool for the prevention and dietary management of hypertension in Nigeria. Patients demonstrated high engagement. Healthcare professionals in Nigeria affirmed the value of care this tool could address. While implementation considerations exist, optimism prevails. Further validation and careful introduction are warranted to facilitate optimal FFQ adoption and impact. Overall, the feasibility outcomes justify progressed research on this FFQ as a practical means to improve dietary assessment practices to prevent and control hypertension better in Nigeria.

## Data Availability

All relevant data are within the manuscript and its Supporting Information files. others shall be made available on request.

## Acknowledgement

We extend our deepest gratitude to the patients and esteemed healthcare professionals of Rivers State University Teaching Hospital, Port Harcourt, Nigeria, whose invaluable participation and insights were instrumental in the successful execution of this study. We would like to extend special thanks to Drs. Nkiru Ahiakwo, Ifeoma Enyoghasim, Eneyo Nelly, Comfort Imarhiagbe, Janny Ikurayeke, Valentine Kogbara, Titi Owen, Dickson Christian, Anwuri Grend, Anita Oweredaba, Ununuma Oguzor, Josephine Sokolo of the Family Medicine Dept RSUTH and Drs. Ibieneiyi, Edith Reuben, Elile Okpara, Imaobong Nonju, Siya George-Batubo, Chinnasa Nzokurum, and Prof. Amah-Tariah for their generous provision of qualitative feedback on the screening tool (FFQ). Their contributions significantly enriched the quality of our research. Furthermore, we express our sincere appreciation to the management of the hospital and the Department of Internal Medicine and Family Medicine for affording us the platform and granting ethical approval, which were pivotal in facilitating the smooth progress of this study.

## Authors’ Contributions

NPB and MAZ collaborated on the research methodology design. NPB led the trial, and NMN and CAA provided technical support. NPB led the analysis and prepared the first draft. MAZ and JBM provided analytical expertise. MAZ, JBM, and CIA provided critical feedback. NPB revised the manuscript. All authors approved the final manuscript.

## Funding

This study was funded by the Tertiary Education Trust Fund (TETFund) of Nigeria. MAZ is currently funded by Wellcome Trust (217446/Z/19/Z). The funders do not have any role in any aspect of this study.

## Conflicts of Interest

None declared.

## Supporting information

**S1 Table.** Food frequency questionnaire

## Notes

### Competing Interest Statement

The authors have declared no competing interest.

### Clinical Protocols

https://doi.org/10.1101/2023.09.25.23296109

### Author Declarations

Ethics committee/IRB of Business, Earth & Environment, Social Sciences (AREA FREC) Committee, University of Leeds, Leeds, United Kingdom gave ethical approval for this work (0484). AND Ethics committee/IRB of Rivers State University Teaching Hospital Research Ethics Committee in Port Harcourt, Nigeria gave ethical approval for this work (RSUTH/REC/2023316).

## References

1. Forouzanfar MH, Liu P, Roth GA, Ng M, Biryukov S, Marczak L, et al. Global Burden of Hypertension and Systolic Blood Pressure of at Least 110 to 115 mm Hg, 1990-2015. JAMA. 2017;317(2):165–82.

2. Dzudie A, Kengne AP, Muna WF, Ba H, Menanga A, Kouam Kouam C, et al. Prevalence, awareness, treatment and control of hypertension in a self-selected sub-Saharan African urban population: a cross-sectional study. BMJ Open. 2012;2(4).

3. Ataklte F, Erqou S, Kaptoge S, Taye B, Echouffo-Tcheugui JB, Kengne AP. Burden of undiagnosed hypertension in sub-saharan Africa: a systematic review and meta-analysis. Hypertension. 2015;65(2):291–8.

4. Odili AN, Chori BS, Danladi B, Nwakile PC, Okoye IC, Abdullahi U, et al. Prevalence, Awareness, Treatment and Control of Hypertension in Nigeria: Data from a Nationwide Survey 2017. Glob Heart. 2020;15(1):47.

5. Akinlua JT, Meakin R, Umar AM, Freemantle N. Current Prevalence Pattern of Hypertension in Nigeria: A Systematic Review. PLoS One. 2015;10(10):e0140021.

6. World Health Organization. The Global Health Observatory (GHO). Prevalence of hypertension among adults aged 30-79 years, age-standardized 2021 [Available from: https://www.who.int/data/gho/data/indicators/indicator-details/GHO/prevalence-of-hypertension-among-adults-aged-30-79-years.

7. Oguoma VM, Nwose EU, Skinner TC, Digban KA, Onyia IC, Richards RS. Prevalence of cardiovascular disease risk factors among a Nigerian adult population: relationship with income level and accessibility to CVD risks screening. BMC Public Health. 2015;15:397.

8. Adeloye D, Owolabi EO, Ojji DB, Auta A, Dewan MT, Olanrewaju TO, et al. Prevalence, awareness, treatment, and control of hypertension in Nigeria in 1995 and 2020: A systematic analysis of current evidence. J Clin Hypertens (Greenwich). 2021;23(5):963–77.

9. Whelton PK, Carey RM, Aronow WS, Casey DE, Jr., Collins KJ, Dennison Himmelfarb C, et al. 2017 ACC/AHA/AAPA/ABC/ACPM/AGS/APhA/ASH/ASPC/NMA/PCNA Guideline for the Prevention, Detection, Evaluation, and Management of High Blood Pressure in Adults: Executive Summary: A Report of the American College of Cardiology/American Heart Association Task Force on Clinical Practice Guidelines. Hypertension. 2018;71(6):1269–324.

10. Zhou B, Perel P, Mensah GA, Ezzati M. Global epidemiology, health burden and effective interventions for elevated blood pressure and hypertension. Nat Rev Cardiol. 2021;18(11):785–802.

11. Zhou B, Bentham J, Di Cesare M, Bixby H, Danaei G, Cowan MJ, … & et al, . Worldwide trends in blood pressure from 1975 to 2015: a pooled analysis of 1479 population-based measurement studies with 19.1 million participants. Lancet. 2017;389(10064):37–55.

12. Batubo NP, Moore JB, Zulyniak MA. Dietary factors and hypertension risk in West Africa: a systematic review and meta-analysis of observational studies. J Hypertens. 2023;41(9):1376–88.

13. Ojji D, Nigeria Sodium Study T. Developing long-term strategies to reduce excess salt consumption in Nigeria. Eur Heart J. 2022;43(13):1277–9.

14. Oyebode O, Oti S, Chen YF, Lilford RJ. Salt intakes in sub-Saharan Africa: a systematic review and meta-regression. Popul Health Metr. 2016;14:1.

15. Mustapha, Mudashiru A, Fakokunde, Olubamiji T, Awolusi, Dele O, editors. The Quick Service Restaurant Business in Nigeria: Exploring the Emerging Opportunity for Entrepreneurial Development and Growth2014.

16. Ogah OS, Okpechi I, Chukwuonye, II, Akinyemi JO, Onwubere BJ, Falase AO, et al. Blood pressure, prevalence of hypertension and hypertension related complications in Nigerian Africans: A review. World J Cardiol. 2012;4(12):327–40.

17. Schmidt BM, Durao S, Toews I, Bavuma CM, Hohlfeld A, Nury E, et al. Screening strategies for hypertension. Cochrane Database Syst Rev. 2020;5(5):CD013212.

18. David L. Streiner, Geoffrey R. Norman, John Cairney. Health measurement scales. A practical guide to their development and use. 5 ed: University Press, Oxford.; 2015. 432 p.

19. Liberati A, Altman DG, Tetzlaff J, Mulrow C, Gotzsche PC, Ioannidis JP, et al. The PRISMA statement for reporting systematic reviews and meta-analyses of studies that evaluate health care interventions: explanation and elaboration. PLoS Med. 2009;6(7):e1000100.

20. Wilson JM, Jungner YG. [Principles and practice of mass screening for disease]. Bol Oficina Sanit Panam. 1968;65(4):281–393.

21. Lee RD, Nieman DC. Nutritional assessment. 6 ed: McGraw-Hill, New York, NY,; 2013.

22. Murphy SP, Wilkens LR, Hankin JH, Foote JA, Monroe KR, Henderson BE, et al. Comparison of two instruments for quantifying intake of vitamin and mineral supplements: a brief questionnaire versus three 24-hour recalls. Am J Epidemiol. 2002;156(7):669–75.

23. Willett WC, Sampson L, Stampfer MJ, Rosner B, Bain C, Witschi J, et al. Reproducibility and validity of a semiquantitative food frequency questionnaire. Am J Epidemiol. 1985;122(1):51–65.

24. Goulet J, Nadeau G, Lapointe A, Lamarche B, Lemieux S. Validity and reproducibility of an interviewer-administered food frequency questionnaire for healthy French-Canadian men and women. Nutr J. 2004;3:13.

25. Bosu WK. An overview of the nutrition transition in West Africa: implications for non-communicable diseases. Proc Nutr Soc. 2015;74(4):466–77.

26. Herman CR, Gill HK, Eng J, Fajardo LL. Screening for preclinical disease: test and disease characteristics. AJR Am J Roentgenol. 2002;179(4):825–31.

27. Paxton AE, Strycker LA, Toobert DJ, Ammerman AS, Glasgow RE. Starting the conversation performance of a brief dietary assessment and intervention tool for health professionals. Am J Prev Med. 2011;40(1):67–71.

28. Cahill E, Schmidt SR, Henry TL, Kumar G, Berney S, Bussey-Jones J, et al. Qualitative research study on addressing barriers to healthy diet among low-income individuals at an urban, safety-net hospital. BMJ Nutr Prev Health. 2020;3(2):383–6.

29. Ball LE, Hughes RM, Leveritt MD. Nutrition in general practice: role and workforce preparation expectations of medical educators. Aust J Prim Health. 2010;16(4):304–10.

30. Chan AW, Tetzlaff JM, Altman DG, Laupacis A, Gotzsche PC, Krleza-Jeric K, et al. SPIRIT 2013 statement: defining standard protocol items for clinical trials. Ann Intern Med. 2013;158(3):200–7.

31. Portz JD, Bayliss EA, Bull S, Boxer RS, Bekelman DB, Gleason K, et al. Using the Technology Acceptance Model to Explore User Experience, Intent to Use, and Use Behavior of a Patient Portal Among Older Adults With Multiple Chronic Conditions: Descriptive Qualitative Study. J Med Internet Res. 2019;21(4):e11604.

32. Federal Ministry of Health. National Nutritional Guideline On Non-Communicable Disease Prevention, Control and Management In: Federal Ministry of Health A, editor. 2014. p. 62.

33. Flesch R. A new readability yardstick. J Appl Psychol. 1948;32(3):221–33.

34. Willett W. Foreword. The validity of dietary assessment methods for use in epidemiologic studies. Br J Nutr. 2009;102 Suppl 1:S1–2.

35. Adebamowo SN, Eseyin O, Yilme S, Adeyemi D, Willett WC, Hu FB, et al. A Mixed-Methods Study on Acceptability, Tolerability, and Substitution of Brown Rice for White Rice to Lower Blood Glucose Levels among Nigerian Adults. Front Nutr. 2017;4:33.

36. Steinemann N, Grize L, Ziesemer K, Kauf P, Probst-Hensch N, Brombach C. Relative validation of a food frequency questionnaire to estimate food intake in an adult population. Food Nutr Res. 2017;61(1):1305193.

37. Mertens E, Kuijsten A, Geleijnse JM, Boshuizen HC, Feskens EJM, Van’t Veer P. FFQ versus repeated 24-h recalls for estimating diet-related environmental impact. Nutr J. 2019;18(1):2.

38. Hennink M, Kaiser BN. Sample sizes for saturation in qualitative research: A systematic review of empirical tests. Soc Sci Med. 2022;292:114523.

39. Thabane L, Ma J, Chu R, Cheng J, Ismaila A, Rios LP, et al. A tutorial on pilot studies: the what, why and how. BMC Med Res Methodol. 2010;10:1.

40. Gupta KK, Attri JP, Singh A, Kaur H, Kaur G. Basic concepts for sample size calculation: Critical step for any clinical trials! Saudi J Anaesth. 2016;10(3):328–31.

41. R Core Team. A language and environment for statistical computing. R Foundation for Statistical Computing. 4.3.1 ed: Vienna, Austria; 2020.

42. Lumivero. QSR International. NVivo (Version 14). NVivo (Version 14) ed2020.

43. Forman J, Damschroder L. Qualitative Content Analysis. Empirical Research for Bioethics: A Primer. Oxford, UK: Elsevier Publishing; 2008.

44. Vaismoradi M, Turunen H, Bondas T. Content analysis and thematic analysis: Implications for conducting a qualitative descriptive study. Nurs Health Sci. 2013;15(3):398–405.

45. Marks GC, Hughes MC, van der Pols JC. Relative validity of food intake estimates using a food frequency questionnaire is associated with sex, age, and other personal characteristics. J Nutr. 2006;136(2):459–65.

46. Subar AF, Potischman N, Dodd KW, Thompson FE, Baer DJ, Schoeller DA, et al. Performance and Feasibility of Recalls Completed Using the Automated Self-Administered 24-Hour Dietary Assessment Tool in Relation to Other Self-Report Tools and Biomarkers in the Interactive Diet and Activity Tracking in AARP (IDATA) Study. J Acad Nutr Diet. 2020;120(11):1805–20.

47. Park Y, Dodd KW, Kipnis V, Thompson FE, Potischman N, Schoeller DA, et al. Comparison of self-reported dietary intakes from the Automated Self-Administered 24-h recall, 4-d food records, and food-frequency questionnaires against recovery biomarkers. Am J Clin Nutr. 2018;107(1):80–93.

48. Kristal AR, Feng Z, Coates RJ, Oberman A, George V. Associations of race/ethnicity, education, and dietary intervention with the validity and reliability of a food frequency questionnaire: the Women’s Health Trial Feasibility Study in Minority Populations. Am J Epidemiol. 1997;146(10):856–69.

49. Hebert JR, Clemow L, Pbert L, Ockene IS, Ockene JK. Social desirability bias in dietary self-report may compromise the validity of dietary intake measures. Int J Epidemiol. 1995;24(2):389–98.

50. Kristal AR, Kolar AS, Fisher JL, Plascak JJ, Stumbo PJ, Weiss R, et al. Evaluation of web-based, self-administered, graphical food frequency questionnaire. J Acad Nutr Diet. 2014;114(4):613–21.

51. Dehghan M, del Cerro S, Zhang X, Cuneo JM, Linetzky B, Diaz R, et al. Validation of a semi-quantitative Food Frequency Questionnaire for Argentinean adults. PLoS One. 2012;7(5):e37958.

52. Illner AK, Freisling H, Boeing H, Huybrechts I, Crispim SP, Slimani N. Review and evaluation of innovative technologies for measuring diet in nutritional epidemiology. Int J Epidemiol. 2012;41(4):1187–203.

53. Oche M, Adamu H. Determinants of patient waiting time in the general outpatient department of a tertiary health institution in north Western Nigeria. Ann Med Health Sci Res. 2013;3(4):588–92.

54. Ajayi IO. Patients’ waiting time at an outpatient clinic in Nigeria--can it be put to better use? Patient Educ Couns. 2002;47(2):121–6.

55. Abah V. O. Hospital Waiting Time, Satisfaction with Services and Patient Arrival Patterns among Primary Care Attendees in a Tertiary Hospital: The Need for Time Specific Appointment Systems. IOSR Journal of Dental and Medical Sciences 2021;20(3):14–27.

56. Jackson KA, Byrne NM, Magarey AM, Hills AP. Minimizing random error in dietary intakes assessed by 24-h recall, in overweight and obese adults. Eur J Clin Nutr. 2008;62(4):537–43.

57. Appel LJ, Moore TJ, Obarzanek E, Vollmer WM, Svetkey LP, Sacks FM, et al. A clinical trial of the effects of dietary patterns on blood pressure. New England journal of medicine. 1997;336(16):1117–24.

58. Sacks FM, Svetkey LP, Vollmer WM, Appel LJ, Bray GA, Harsha D, et al. Effects on blood pressure of reduced dietary sodium and the Dietary Approaches to Stop Hypertension (DASH) diet. New England journal of medicine. 2001;344(1):3–10.

59. Filippou CD, Tsioufis CP, Thomopoulos CG, Mihas CC, Dimitriadis KS, Sotiropoulou LI, et al. Dietary Approaches to Stop Hypertension (DASH) Diet and Blood Pressure Reduction in Adults with and without Hypertension: A Systematic Review and Meta-Analysis of Randomized Controlled Trials. Adv Nutr. 2020;11(5):1150–60.

60. Bharati R, Kovach KA, Bonnet JP, Sayess P, Polk E, Harvey K, et al. Incorporating Lifestyle Medicine Into Primary Care Practice: Perceptions and Practices of Family Physicians. Am J Lifestyle Med. 2023;17(5):704–16.

61. Clarke CA, Hauser ME. Lifestyle Medicine: A Primary Care Perspective. J Grad Med Educ. 2016;8(5):665–7.

62. Joseph RP, Daniel CL, Thind H, Benitez TJ, Pekmezi D. Applying Psychological Theories to Promote Long-Term Maintenance of Health Behaviors. Am J Lifestyle Med. 2016;10(6):356–68.

63. Levine DM, Green LW, Deeds SG, Chwalow J, Russell RP, Finlay J. Health education for hypertensive patients. JAMA. 1979;241(16):1700–3.

64. Lianov L, Johnson M. Physician competencies for prescribing lifestyle medicine. JAMA. 2010;304(2):202–3.

65. Kruger R, Stonehouse W, von Hurst PR, Coad J. Combining food records with in-depth probing interviews improves quality of dietary intake reporting in a group of South Asian women. Aust N Z J Public Health. 2012;36(2):135–40.

66. Blumenthal JA, Babyak MA, Sherwood A, Craighead L, Lin PH, Johnson J, et al. Effects of the dietary approaches to stop hypertension diet alone and in combination with exercise and caloric restriction on insulin sensitivity and lipids. Hypertension. 2010;55(5):1199–205.

67. Bricarello LP, de Almeida Alves M, Retondario A, de Moura Souza A, de Vasconcelos FAG. DASH diet (Dietary Approaches to Stop Hypertension) and overweight/obesity in adolescents: The ERICA study. Clin Nutr ESPEN. 2021;42:173–9.

68. Sedgwick P. Cross sectional studies: advantages and disadvantages. BMJ : British Medical Journal. 2014;348:g2276.

69. Hu FB, Stampfer MJ, Rimm E, Ascherio A, Rosner BA, Spiegelman D, et al. Dietary fat and coronary heart disease: a comparison of approaches for adjusting for total energy intake and modeling repeated dietary measurements. Am J Epidemiol. 1999;149(6):531–40.

70. Svetkey LP, Simons-Morton DG, Proschan MA, Sacks FM, Conlin PR, Harsha D, et al. Effect of the dietary approaches to stop hypertension diet and reduced sodium intake on blood pressure control. J Clin Hypertens (Greenwich). 2004;6(7):373–81.

71. Soguel L, Vaucher C, Bengough T, Burnand B, Desroches S. Knowledge Translation and Evidence-Based Practice: A Qualitative Study on Clinical Dietitians’ Perceptions and Practices in Switzerland. J Acad Nutr Diet. 2019;119(11):1882–9.

72. Moutou KE, England C, Gutteridge C, Toumpakari Z, McArdle PD, Papadaki A. Exploring dietitians’ practice and views of giving advice on dietary patterns to patients with type 2 diabetes mellitus: A qualitative study. J Hum Nutr Diet. 2022;35(1):179–90.

73. Baranowski T, Willett W. 4924-Hour Recall and Diet Record Methods. Nutritional Epidemiology: Oxford University Press; 2012. p. 0.

74. Ammerman AS, DeVellis RF, Carey TS, Keyserling TC, Strogatz DS, Haines PS, et al. Physician-based diet counseling for cholesterol reduction: current practices, determinants, and strategies for improvement. Prev Med. 1993;22(1):96–109.

75. Damschroder LJ, Aron DC, Keith RE, Kirsh SR, Alexander JA, Lowery JC. Fostering implementation of health services research findings into practice: a consolidated framework for advancing implementation science. Implement Sci. 2009;4:50.

76. Lemon SC, Zapka J, Li W, Estabrook B, Rosal M, Magner R, et al. Step ahead a worksite obesity prevention trial among hospital employees. American journal of preventive medicine. 2010;38(1):27–38.

77. Peters DH, Adam T, Alonge O, Agyepong IA, Tran N. Implementation research: what it is and how to do it. BMJ. 2013;347:f6753.

